# Persistence of child marriage in rural Bangladesh and impact on maternal and perinatal health: findings from a health and demographic surveillance system

**DOI:** 10.1101/2021.09.06.21263190

**Authors:** Kyu Han Lee, Atique I. Chowdhury, Qazi S. Rahman, Solveig A. Cunningham, Shahana Parveen, Sanwarul Bari, Shams E. Arifeen, Emily S. Gurley

## Abstract

**Objectives:** To describe temporal trends in child marriage between 1990 and 2019 in a rural sub-district of Bangladesh and characterize relationships between age, time to pregnancy, complications during delivery, and perinatal mortality.

**Design:** Health and demographic surveillance system.

**Setting:** Baliakandi sub-district, Bangladesh.

**Participants:** 56,155 female residents.

**Main outcome measures:** Annual proportion of marriages to female residents under 18 years of age, time between first marriage and pregnancy, proportion of births with complications during delivery, and odds of perinatal mortality.

**Results:** Between 1990 and 2010, the proportion of marriages to female residents under 18 years of age decreased from 71% to 57%. Most notably, marriages to females aged 10 to 12 years dropped from 22% of all marriages to 3%. In 2019, 53% of all marriages were to females under 18 years. The estimated time between first marriage and pregnancy did not differ by female age at marriage. By 365 days after marriage, the cumulative incidence of pregnancy was approximately 50% for each age group. Adolescent girls were more likely to experience complications during delivery with at least one complication reported for 36% of mothers aged 13 to 15 years, 32% of mothers aged 16 to 17 years, and 23% of mothers aged 18 to 34 years (χ^2^ test, P<0.001). Compared to adults, births among females aged 13 to 15 years were more likely to result in stillbirths (odds ratio 2.23, 95% confidence interval 1.19 to 4.16) and births among females 16-17 years were more likely to result in early neonatal deaths (odds ratio 1.57, 95% confidence interval 1.01 to 2.42).

**Conclusions:** Child marriage persists in Baliakandi. Over half of all marriages were to child brides and only minor reductions were seen over the past decade. Pregnancies were common among adolescent girls with no evidence of delayed pregnancy after marriage. Compared to adults, adolescents were more likely to experience complications during delivery and perinatal death. Preventing child marriage has substantial social and health benefits for girls and, by doing so, will also contribute to Bangladesh’s commitment to reduce child mortality.

## INTRODUCTION

In 2019 alone, approximately 5.2 million children under-five died worldwide[1]. While remarkable reductions in mortality have been observed over the past two decades among older children, progress has been slow in reducing neonatal deaths, which now account for nearly half of all under-five mortality [1]. Even less progress has been made in preventing stillbirths. Globally, an estimated 2 million babies are stillborn at 28 weeks or more of gestation each year [2]. Current trends in under-five mortality and stillbirths suggest we will not achieve our global targets set for 2030, which includes the Sustainable Development Goal (SDG) target of 25 under-five deaths per 1,000 live births [1] and the Every Newborn Action Plan target of 12 stillbirths per 1,000 total births [2].

Girls who become pregnant during adolescence are at increased risk for experiencing a perinatal death [3] and for obstetric complications including pregnancy-induced hypertension, obstructed labor, obstetric fistula, and postpartum hemorrhage [4–7]. These complications during pregnancy and childbirth are the leading cause of death among adolescent girls [8]. These risks are exacerbated by the persistent practice of child marriage [9], a human rights violation that profoundly impacts one of the most vulnerable populations: adolescent girls. Child marriage, defined as the formal or informal union of a child under 18 years of age [10], disproportionately affects girls, particularly in low- and middle-income countries [11], and contributes to severe social, developmental, and reproductive harms [12]. Although extensive global efforts have been made to end child marriage, progress has been stagnant in recent years. In 2018, 21% of women aged 20-24 years worldwide were married before 18 years of age and, at the current rate, it would take at least 50 years to end child marriage, decades past yet another SDG target [11].

Globally, an estimated 16 million girls aged 15 to 19 become pregnant each year and 90% of these adolescent pregnancies occur within marriage [9]. Given societal and familial pressures to bear children immediately after marriage in many communities [12], marriage often means the beginning of a sexual relationship for children who are still in the process of maturing both physically and cognitively [3].

Some of the highest rates of child marriage are found in Bangladesh. In 2018, 59% of women aged 20 to 24 years were married before 18 years of age and 28% of married adolescents aged 15 to 19 reported a pregnancy [13]. In 2017, the Child Health and Mortality Prevention Surveillance (CHAMPS) network established a health and demographic surveillance system (HDSS) in Baliakandi, a rural sub-district of Bangladesh with approximately 216,000 residents [14]. We used the Baliakandi HDSS to characterize temporal trends in child marriage in this rural region of Bangladesh. We examined whether pregnancy was delayed for newly married adolescents and examined the relationship between maternal age, complications during delivery, and perinatal mortality.

## METHODS

### Health and demographic surveillance system

We used data from an ongoing HDSS established in Baliakandi, Bangladesh. Baseline surveys were conducted between March and August 2017 during which household and sociodemographic data were collected from all residents. Married females under 50 years of age completed a birth history questionnaire either during baseline surveys or after in-migration.

Four to six data rounds were conducted every year between September 2017 and February 2020, during which we identified key demographic events including marriages, pregnancies, pregnancy outcomes, deaths, and out-migrations. As missed demographic events can be identified in subsequent data rounds, we used HDSS data as of February 2020 for events that occurred between September 2017 and August 2019 for most analyses. Each year, we observed consent rates at or greater than 99% among approximately 52,000 households. Given regular follow up, missing data were uncommon for variables included in this analysis. Individuals with missing data were excluded from relevant analysis and the total missing was noted in the results. Additional details on the Baliakandi HDSS are described by Cunningham et al [14].

We defined stillbirth as fetal demise at or after 28 weeks of estimated gestational age and early neonatal death as death within the first 7 days after live birth. Stillbirths and early neonatal deaths were considered perinatal deaths.

### Age at marriage among female residents

We estimated the annual proportion of marriages to children among female residents married between 1990 and 2019 using data from birth history questionnaires and marriage events identified data rounds between September 2017 and December 2019 (Supplementary figure 1). The age difference between partners was calculated among first marriages that occurred in Baliakandi between September 2017 and December 2019.

### Time between first marriage and childbearing

We estimated time between first marriage and pregnancy among female residents under 35 years of age who remained in Baliakandi for at least 180 days after marriage (Supplementary figure 1). Females who in-migrated after marriage were excluded from the analysis as marriage dates were limited only to the month and year and dates were collected retrospectively, which could introduce systematic bias through misreporting [15]. The last menstrual period was used as a proxy of pregnancy date. We used Kaplan-Meier estimates to examine the relationship between the age at first marriage and time between marriage and pregnancy. Age was categorized as 12-15 years, 16-17 years, and 18-34 years. To explore potential bias introduced by out-migration, we compared the age of female residents who exited Baliakandi within 180 days of first marriage and those who remained for at least 180 days after first marriage. We further restricted survival analysis to female residents who remained in Baliakandi for at least 365 days after marriage.

### Maternal age at birth and perinatal death

We used logistic regression to estimate the association between maternal age at birth and perinatal death among all singleton births that occurred between September 2017 and August 2019. We used multinomial logistic regression to estimate the association between maternal age at birth and stillbirths and early neonatal deaths (Supplementary figure 2). Age was categorized as 13-15 years, 16-17 years, and 18-34 years. We repeated the model after adjusting for parity and household wealth. Household wealth was categorized as a quintile based on the Demographic and Health Survey (DHS) wealth index score [16].

### Complications during delivery

In December 2018, we supplemented the Baliakandi HDSS with detailed pregnancy surveillance (Supplementary figure 2). All pregnant women identified during data rounds were contacted within 3 days of giving birth to collect information on complications during delivery. We estimated the frequency of complications during delivery by maternal age at birth for all pregnancy outcomes between January and August 2019. For live births, we estimated the frequency of preterm births, defined as birth before an estimated gestational age of 37 weeks. Gestational age was estimated using the last menstrual period. Chi-squared test and Fisher’s exact test were used to test for statistical differences by maternal age.

## RESULTS

### Age at marriages among female residents

A total 56,155 female residents of Baliakandi were married between 1990 and 2019 and all had age at marriage. Among female residents married in 1990, 71% were married before 18 years of age (915/1,284). This proportion dropped to 57% among female residents married in 2010 (1,185/2,063). Between 1990 and 2010, the proportion of marriages to females under 13 years decreased from 22% (288/1,284) to 3% (72/2,063), females aged 13 to 15 years decreased from 32% (411/1,284) to 26% (538/2,063) and females aged 16 to 17 years increased from 17% (216/1,284) to 28% (575/2,063) (Figure 1). The mean age at marriage increased from 15.3 years (standard deviation (SD) 3.2) to 17.3 years (SD 3.4).

**Figure 1.**
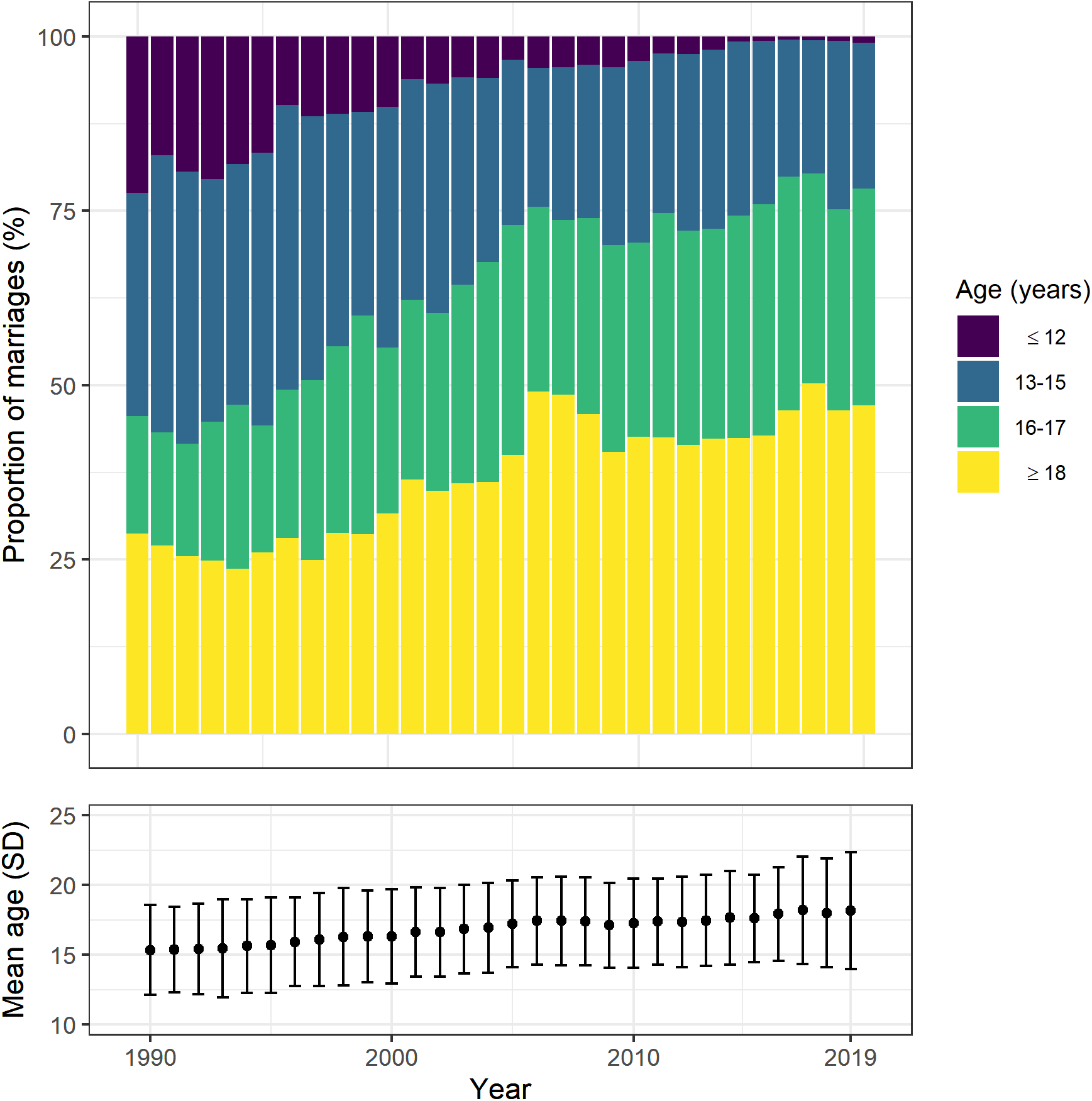
Age at marriage among 56,155 female residents of Baliakandi sub-district, Bangladesh, 1990 to 2019. Bars represent the annual proportion of marriages to specific age groups. Points and error bars represent the mean age at marriage and standard deviation.

Between 2010 and 2019, the overall proportion of marriages to female residents under 18 years dropped from 57% (1,185/2,063) to 53% (1,698/3,210). The proportion to females under 13 years decreased from 3% (72/2,063) to 1% (28/3,210), females aged 13 to 15 years decreased from 26% (538/2,063) to 21% (671/3,210), females aged 16 to 17 years increased from 28% (575/2,063) to 31% (999/3,210). The mean age at marriage increased from 17.3 years (SD 3.4) to 18.2 years (SD 4.2).

A total 3,860 first marriages were identified in Baliakandi between September 2017 and August 2019. Among all first marriages, males were a median of 8 years (interquartile range (IQR) 5 to 10) older than their female spouses. The median age difference was 9.5 years (IQR 8 to 12) for females married before 13 years of age, 9 years (IQR 6 to 12) for those married at ages13 to 15 years, 8 years (IQR 6 to 10) for those married at ages 16 to 17 years, and 6 years (IQR 3 to 9) among those married at ages 18 to 34 years. We excluded one female resident with missing spouse age.

### First marriages and out-migration

A total 3,764 female residents under 35 years of age reported first marriages in Baliakandi between September 2017 and August 2019 (Supplementary table 1). Younger brides were more likely to be in the lowest household wealth quintile: 25% among ages 10 to 15 years (271/1,077), 20% among ages 16-17 years (227/1,150), and 15% among females ages 18 to 34 years (236/1,537) (χ^2^ test, P<0.001). We excluded one resident with missing household wealth data.

Out-migration was common among newly married females with 51% (1,940/3,764) exiting Baliakandi within the first 180 days of marriage (Supplementary table 1). This proportion included females with less than 180 days of follow up by end of August 2019. Most out-migrations occurred immediately after marriage with 45% of newly married women (1,694/3,764) out-migrated within a week of marriage. Out-migrants were less likely to be child brides (1,090/1,940, 56%) compared to those who stayed in Baliakandi (1137/1,824, 62%; χ^2^ test, P<0.001).

### Time between first marriage and pregnancy

As pregnancies could not be captured among out-migrants, we estimated time between first marriage and pregnancy for 1,320 female residents who were newly married in Baliakandi between September 2017 and August 2019 and remained in Baliakandi for at least 180 days after marriage. Fifty-four percent (715/1,320) reported at least one pregnancy by August 2019. This proportion includes females with less than 180 days of follow up after marriage. The mean time to pregnancy or end of follow up was 264 days. Thirty-three percent (239/715) of first pregnancies were among females married at ages 12 to 15 years, 29% (209/715) were among those married at ages 16 to 17 years and 37% (267/715) were among those married at ages 18 to 34 years.

Time between marriage and pregnancy was estimated using the last menstrual period. We estimated Kaplan-Meier cumulative incidence curves stratified by age at marriage and did not find a statistically significant difference by age in the timing of pregnancy (log-rank test, P=0.535) (Figure 2). Within 365 days of marriage, approximately half of all newly married females had a pregnancy: 52% (95% confidence interval (CI) 47 to 57%) among those married at 12 to 15 years, 53% (95% CI 48 to 59%) among those married at ages 16 to 17 years and 50% (95% CI 45 to 54%) among those married at ages 18 to 34 years. Results were similar after restricting the analysis to female residents who remained in Baliakandi for at least 365 days after marriage (supplementary table 1).

**Figure 2.**
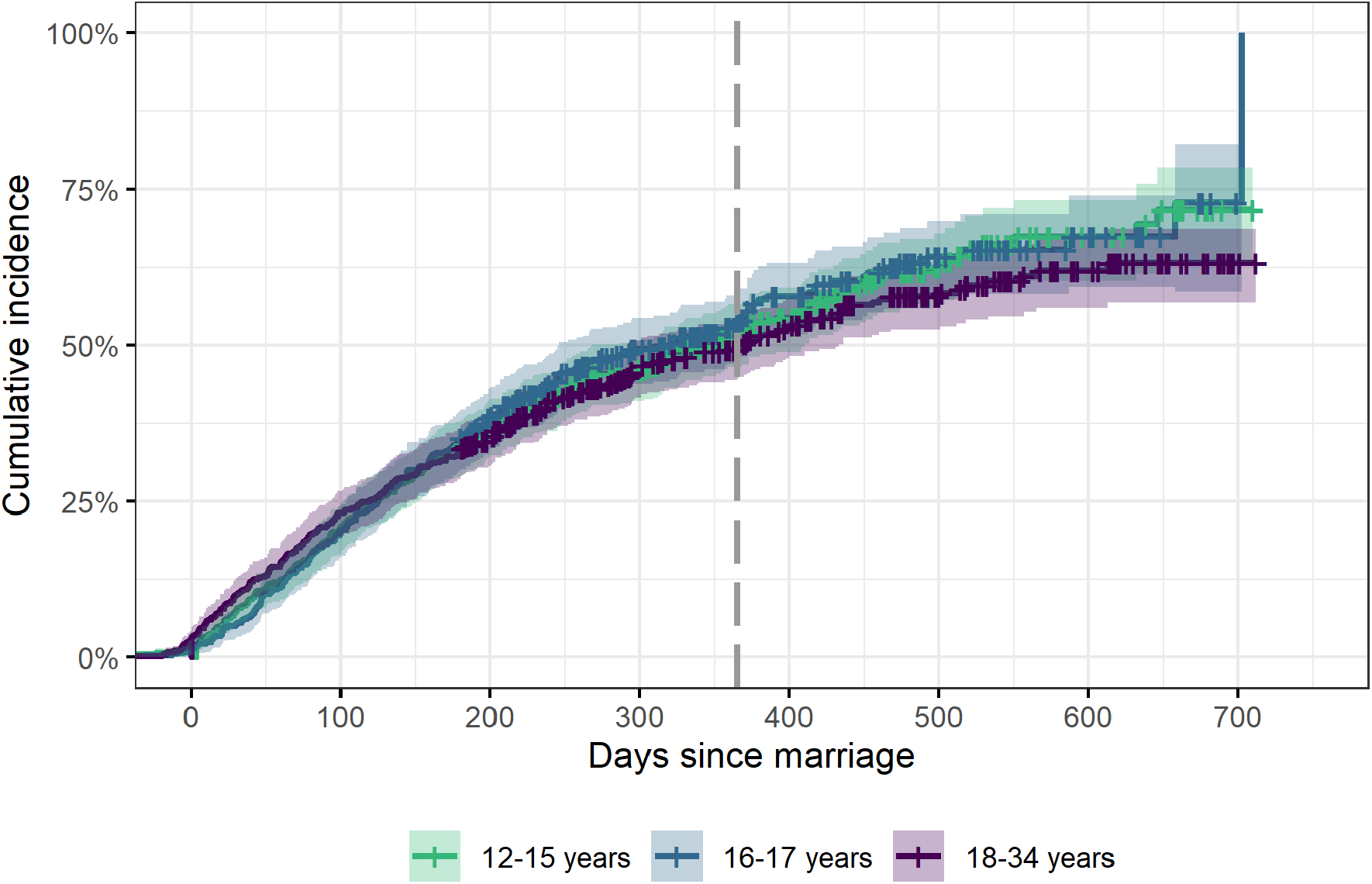
Cumulative incidence of pregnancies after first marriage and 95% confidence intervals, by age at marriage. Among 1,320 females under 35 years of age who remained in Baliakandi for at least 180 days after marriage, Baliakandi sub-district, Bangladesh. Grey dashed line indicates 365 days after marriage.

### Pregnancy outcomes and perinatal deaths

We identified a total 10,345 pregnancy outcomes in Baliakandi between September 2017 and August 2019: 269 (3%) among females aged 12 to 15 years, 863 (8%) among females aged 16 to 17 years and 9,213 (89%) among females aged 18 years or older. Pregnancy outcomes included 1,018 (10%) miscarriages/abortions, 217 (2%) singleton stillbirths, and 9,024 (87%) singleton live births. Outcomes of multiple gestations included 75 (1%) twin live births, 10 (0%) twin births with at least 1 stillbirth, and 1 (0%) triplet live birth.

We identified a total 446 perinatal deaths. Two hundred thirty (52%) were stillbirths, 96 (22%) were neonatal deaths that occurred within 24 hours after birth, and 120 (27%) were early neonatal deaths that occurred after 24 hours (Figure 3). Nineteen (4%) perinatal deaths were among females aged 13 to 15 years, 48 (11%) were among females aged 16 to 17 years, 282 (63%) were among females aged 18 to 29 years, and 97 (22%) were among females aged 30 years and above. There were no perinatal deaths among females under 13 years of age. This resulted in perinatal mortality rates of 79 perinatal deaths per 1,000 total births among females aged 13 to 15 years, 62 perinatal deaths per 1,000 total births among females aged 16 to 17, 42 perinatal deaths per 1,000 total births among females aged 18 to 29 years, and 58 perinatal deaths per 1,000 total births among females aged 30 years and above.

**Figure 3.**
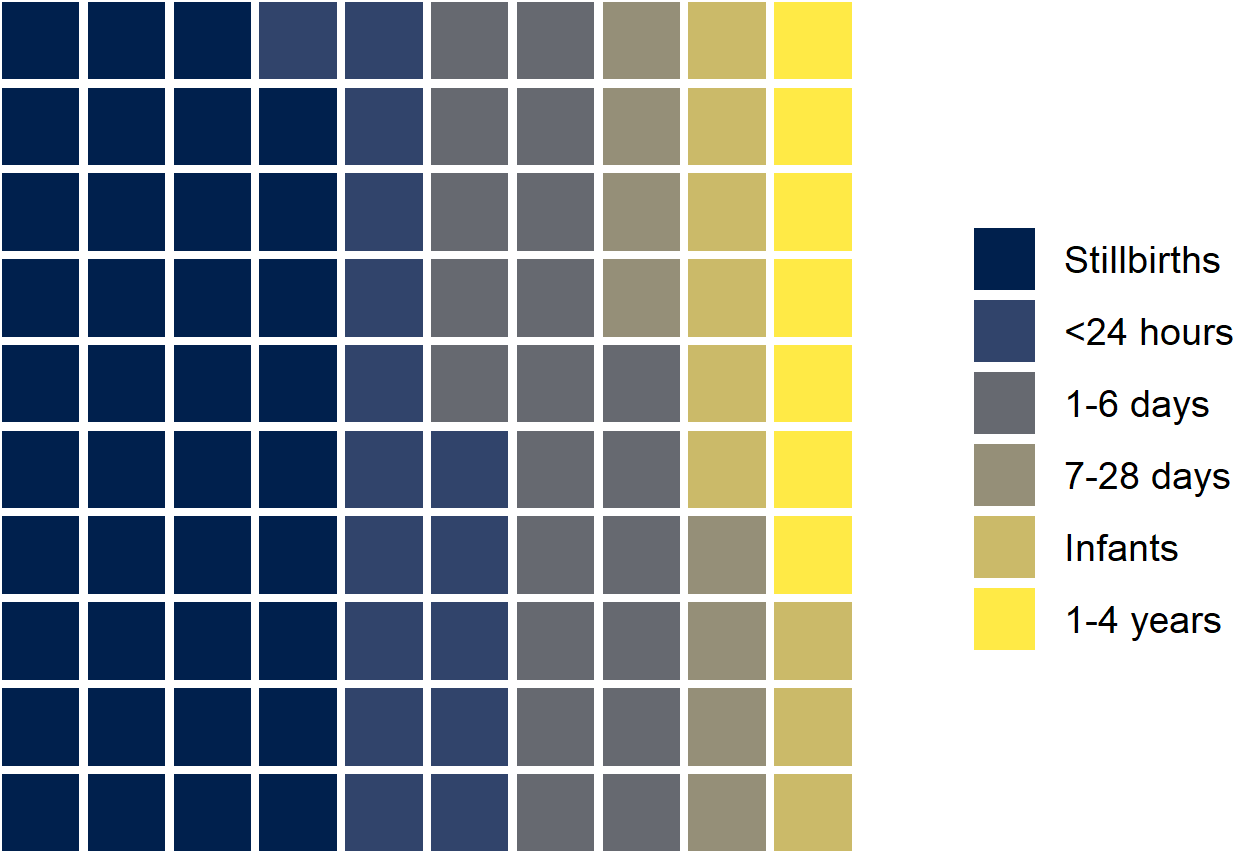
Proportion of stillbirths and early neonatal deaths among 446 perinatal deaths, September 2017 to August 2019, Baliakandi sub-district, Bangladesh. Each square represents one percent of the total.

### Maternal age at birth and perinatal mortality among singleton births

To examine the relationship between maternal age at birth and perinatal mortality, we restricted our analysis to singleton births among female residents under 35 years of age (Table 1). Among singleton births identified between September 2017 and August 2019, 2% (198/8,806) were stillbirths and 2% (188/8,806) ended in early neonatal deaths. The estimated stillbirth rate was 22 stillbirths per 1,000 singleton births and the estimated early neonatal mortality rate was 22 deaths per 1,000 singleton live births. Eighty-four percent of perinatal deaths were among females under 30 years of age (324/386) and 52% were among first-time mothers (202/386). Fifty-one percent were among first-time mothers under 30 years of age (198/386).

**Table 1.** Characteristics of singleton births among females under 35 years of age, Baliakandi sub-district, Bangladesh, September 2017 to August 2019.

| Characteristic | All births<br>N=8,806<br>n (%) | Live births<br>surviving 7 days<br>N=8,420<br>n (%) | Stillbirths<br>N=198<br>n (%) | Early neonatal deaths<br>N=188<br>n (%) |
| --- | --- | --- | --- | --- |
| Maternal age at delivery (years)* |  |  |  |  |
| 12 to 15 | 238 (3%) | 221 (3%) | 11 (6%)<br>Rate: 46 per 1,000<br>births | 6 (3%)<br>Rate: 26 per 1,000 live<br>births |
| 16 to 17 | 769 (9%) | 725 (9%) | 20 (10%)<br>Rate: 26 per 1,000<br>births | 24 (13%)<br>32 per 1,000 live births |
| 18 to 34 | 7,799 (89%) | 7,474 (89%) | 167 (84%)<br>Rate: 21 per 1,000<br>births | 158 (84%)<br>21 per 1,000 live births |
| Parity |  |  |  |  |
| Nulliparous | 3,827 (43%) | 3,625 (43%) | 103 (52%) | 99 (53%) |
| 1 | 3,252 (37%) | 3,146 (37%) | 57 (29%) | 49 (26%) |
| 2 | 1,315 (15%) | 1,261 (15%) | 26 (13%) | 28 (15%) |
| 3+ | 412 (5%) | 388 (5%) | 12 (6%) | 12 (6%) |
| Household wealth quintile |  |  |  |  |
| Highest | 1,815 (21%) | 1,751 (21%) | 28 (14%) | 36 (20%) |
| High | 1,863 (21%) | 1,786 (21%) | 45 (23%) | 32 (22%) |
| Middle | 1,809 (21%) | 1,727 (21%) | 42 (21%) | 40 (21%) |
| Low | 1,689 (19%) | 1,610 (19%) | 37 (19%) | 42 (17%) |
| Lowest | 1,630 (19%) | 1,546 (18%) | 46 (23%) | 38 (19%) |

\*No singleton births were reported among mothers under 12 years of age. Births among women over 34 years were excluded.

Compared to females aged 18 to 34 years, the odds of perinatal death were higher for females aged 13 to 15 years (odds ratio (OR) 1.77, 95% CI 1.07 to 2.93) and females aged 16 to 17 years (OR 1.40, 95% CI 1.01 to 1.93) (Figure 4). When we considered stillbirths and early neonatal deaths as separate outcomes, females aged 13 to 15 years had higher odds of stillbirth (OR 2.23, 95% CI 1.19 to 4.16) and females aged 16 to 17 years had higher odds of early neonatal death (OR 1:57, 95% CI 1.01 to 2.42), compared to females aged 18 to 34 years (Figure 4). Associations were no longer statistically significant for stillbirths (adjusted odds ratio (aOR) 1.77, 95% CI 0.92 to 3.38) and early neonatal deaths (aOR 1.28, 95% CI 0.80 to 2.04), after adjusting for household wealth and parity (Supplementary table 2).

**Figure 4.**
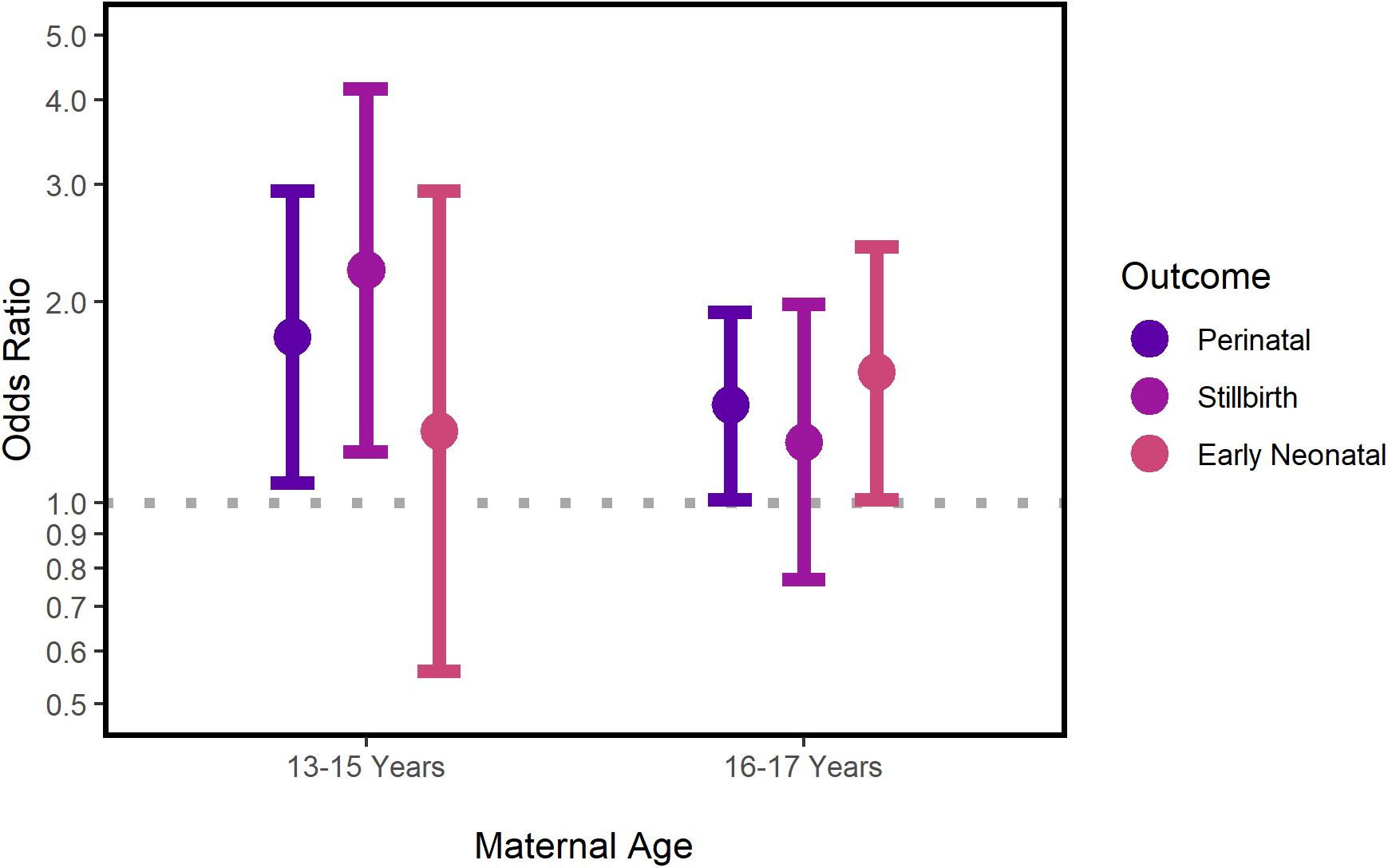
Odds ratios of perinatal death, stillbirth, and early neonatal death compared to births among female residents aged 18 to 34 years. Total 8,806 singleton births among females under 35 years at delivery, Baliakandi sub-district, Bangladesh, September 2017 to August 2019.

### Complications during delivery

Information on complications during delivery were collected through detailed pregnancy surveillance for 3,286 singleton births of a total 3,311 singleton births (99%) that occurred between January and August 2019 among female residents who were under 35 years of age at birth. Complications were common in Baliakandi regardless of maternal age, with 24% of all births (803/3,286) reporting at least one complication, including prolonged/obstructed labor, birth trauma or difficult delivery, and heavy bleeding during delivery (Table 2). Nineteen percent (610/3,286) involved unplanned hospital admissions. Nineteen percent all live births (604/3,223) were considered preterm based on estimated gestational age and 38% (180/604) of preterm births involved pre-labor cesarean sections.

**Table 2.** Complications during delivery among 3,286 singleton births by female residents under 35 years of age, by maternal age at birth, Baliakandi sub-district, Bangladesh, January to August 2019.

| Events | All<br>N=3,286<br>n (%) | 13 to 15*<br>years<br>N=101<br>n (%) | 16 to 17<br>years<br>N=291<br>n (%) | 18 to 34<br>years<br>N=2,894<br>n (%) | P-value<sup>†</sup> |
| --- | --- | --- | --- | --- | --- |
| Any complications<sup>‡</sup> | 803 (24%) | 36 (36%) | 93 (32%) | 674 (23%) | <0.001 |
| Prolonged/obstructed labor | 267 (8%) | 14 (14%) | 33 (11%) | 220 (8%) | 0.008 |
| Birth trauma or difficult delivery | 276 (8%) | 17 (17%) | 42 (14%) | 217 (8%) | <0.001 |
| High blood pressure | 145 (5%) | 4 (4%) | 15 (5%) | 126 (5%) | 0.728 |
| Heavy bleeding during delivery | 120 (4%) | 9 (9%) | 10 (3%) | 101 (4%) | 0.029 |
| Severe headaches with blurred visions | 114 (3%) | 5 (5%) | 11 (4%) | 98 (3%) | 0.550 |
| Fetal malpresentation | 39 (1%) | 2 (2%) | 2 (1%) | 35 (1%) | 0.456 |
| High fever with abdominal pain | 29 (1%) | 1 (1%) | 1 (0%) | 27 (1%) | 0.548 |
| High fever with smelly discharge | 19 (1%) | 1 (1%) | 0 (0%) | 18 (1%) | 0.297 |
| Unplanned hospital admission for delivery | 610 (19%) | 28 (28%) | 79 (27%) | 503 (17%) | <0.001 |
| Preterm birth* | 604/3,223<br>(19%) | 14/98<br>(14%) | 51/287<br>(18%) | 539/2838<br>(19%) | 0.456 |

\*No singleton birth among children under 13 years of age with data on complications during delivery.
<sup>†</sup>Chi-squared test or Fisher's exact test; comparing all three age groups
<sup>‡</sup>Includes any of the listed complications
'Only among live births

Compared to females aged 18 to 34 years, adolescents were more likely to experience at least one complication during delivery (χ^2^ test, P<0.001) and have unplanned admissions (χ^2^ test, P<0.001). Among specific complications, obstructed or prolonged labor, birth trauma or difficult delivery, and heavy bleeding during delivery were more common among females under 18 years of age, compared to those aged 18 to 34 years (χ^2^ test or Fisher’s exact tests, all P<0.050).

## DISCUSSION

Using data from a HDSS covering approximately 216,000 residents in a rural sub-district of Bangladesh, we found almost no change in the proportion of marriages to child brides over the past decade. In 2019, more than half of all marriages were still to child brides. However, we note substantial increases in the age of child brides. We found no evidence of postponed pregnancies for child brides. Half of all newly married females became pregnant within a year of marriage, including adolescents.

Complications during delivery were common in Baliakandi regardless of maternal age. However, they were more likely to occur in adolescent pregnancies. Over a quarter of deliveries among adolescent girls involved an unplanned hospital admission and around a third reported complications, including those related to physical immaturity [3]. Common complications included prolonged or obstructed labor, birth trauma or difficult deliveries, and heavy bleeding, which can lead to severe, long-term urologic, gynecologic and neurologic injuries as well as secondary infertility [17–19].

Adolescent pregnancies were more likely to end in perinatal death than pregnancies among adult women. Risk of perinatal death could partially be explained by parity and household wealth. Adolescent mothers were more likely to be nulliparous, which is associated with increased risk of obstetric complications and perinatal death in Bangladesh [20,21]. Further, less wealthy families were more likely to have child brides as well as perinatal deaths, perhaps due to poor nutrition, lack of resources, and less access of health care [22–25].

### Strengths and weaknesses of this study

A notable strength of this study was the use of HDSS data to investigate child marriage. Most quantitative studies of child marriage in South Asia are based on Demographic and Health Surveys [26,27] or other cross-sectional surveys [28]. Unlike cross-sectional samples, the Baliakandi HDSS prospectively collected key information such as dates of birth, marriage, pregnancies, and births through frequent data rounds, leading to more accurate reporting and estimates. In Matlab, Bangladesh, which hosts a similar HDSS originally established in 1966 [15], almost two-thirds of female residents aged 15 to 29 years misreported their age at first marriage in cross-sectional surveys, with the average reported age nearly 2 years younger than true values in the HDSS [29]. While the exact causes of underreporting were unknown, it may have been driven by a desire to reduce dowry costs, which increase with the bride’s age, and to meet the “preferred age” of brides, which is typically under 20 years [29,30]. If similar underreporting is true for Baliakandi, we may have underestimated child marriage in years prior to 2017 given marriages dates were collected retrospectively for this period. Our estimates did not include female residents who out-migrated nor those who died during this period. While it is unclear how these deaths might impact our estimates, out-migration may have led to underestimates. Our findings from 2017 to 2019 suggest newly married brides who out-migrate were more likely to be child brides than those we remained in Baliakandi. Lastly, our estimates of time between first marriage and pregnancy may have been influenced by downward bias as adult brides were more likely to out-migrate soon after marriage.

### Comparison with other studies

Our findings from Baliakandi were consistent with larger national trends described in the Bangladesh DHS [13]; child marriage remains a pervasive and persistent human rights violation in Bangladesh and little progress has been made in recent years. Over half of all marriages still involve child brides [13]. Associations between adolescent pregnancies and perinatal death were consistent with earlier studies which used the Bangladesh DHS. These cross-sectional studies showed adolescent pregnancies were associated with increased risk of stillbirths and neonatal mortality [21,31]. Associations between maternal age and obstetric complications were consistent with a prior study conducted in northwest Bangladesh [20].

The preterm birth rate in Baliakandi was consistent with national estimates, which is one of the highest in the world [32]. One potential explanation was the common practice of elective cesarean sections in Baliakandi, which may be driven by increased practice without medical indication, especially among private facilities, by a fear of vaginal deliveries, and the belief that cesarean deliveries are safer for the mother and baby [33,34].

### Policy implications and conclusions

Bangladesh has one of highest rates of child marriage in the world [13], even with legislation prohibiting child marriage in existence since 1929 [35]. Critics argue progress may be further hindered by recent legal exceptions introduced in the 2017 Child Marriage Restraint Act, which permits child marriage under ambiguous circumstances that are “in the best interests of the minor” [36].

Globally, 90% of adolescent pregnancies occur within marriage [9]. Although childbearing should be delayed among adolescent girls to protect the health of the mother and newborn [3,12,37], our study showed no evidence of postponed pregnancy among child brides in Baliakandi. Adolescent pregnancies are likely driven by the social value placed on childbearing for newly married wives in Bangladesh and the prospect of improving one’s status within the husband’s family through childbirth [38,39]. Many newly married adolescents lack power and social status to negotiate delays in pregnancy, coupled with a lack of knowledge or misconceptions of contraceptives. One example of misconceptions is that contraceptives can lead to infertility [39]. In contrast, wives who delay childbearing often experience stigma of perceived infertility, abuse by in-laws, and rumors of infidelity [38]. These dynamics may drive low contraceptive use and unmet family planning needs seen among adolescent pregnancies [13]. Adolescent pregnancies are more likely to experience obstetric complications such as prolonged and obstructed labor, which can cause long-term physical consequences [17–19] as well as devastating psychosocial disabilities even after treatment, as girls deal with shame, stigma, and rejection by their families and communities [17].

The problem of child marriage is daunting; it is a practice deeply rooted in social and gender norms and sustained by gender inequality, poverty and insecurity [12,40–42]. However, encouraging commitments have been made to end child marriage in Bangladesh, such as the UNFPA-UNICEF Global Programme to End Child Marriage, which involves targeted actions in 12 focus countries with high burdens of child marriage, including Bangladesh [43]. Recent evaluations of the Global Programme show all global targets have been met or surpassed in the first phase (2016 to 2019). Targets include developing life skills and comprehensive sexuality education among adolescent girls, supporting continued formal or non-formal education among adolescent girls, increasing awareness of the benefits of preventing child marriage in communities, supporting health and protection systems, developing national plans of action, and generating data to accelerate ending child marriage [44]. Further, findings from large randomized control trials such as BALIKA in Bangladesh suggest various interventions such as education support, gender rights awareness training, and livelihood training can lead to large reductions in child marriage. Villages assigned to one of the three aforementioned interventions showed a 25 to 30% reduction in risk of child marriage compared to control villages [45].

Only by accelerating ongoing commitments and promoting additional widespread interventions we can truly eliminate child marriage within the next decade. By addressing child marriage, we improve the health of two groups of children: adolescent mothers and their newborns. We can prevent a chain of harmful social, developmental, and reproductive harms that have lasting impacts on adolescent girls while mitigating the global burden of stillbirths and neonatal deaths.

## Data Availability

Requests for sharing of the data will be considered and reviewed by the study principal investigators. Requests can be made to the corresponding author.

## Ethical approval

The study was approved by the icddr,b human subjects review committee. Written consent was obtained from study participants.

