## Supplementary table 3 for "Persistence of child marriage in rural Bangladesh and impact on maternal and perinatal health: findings from a health and demographic surveillance system"

### Supplementary table 3. Adjusted odds ratios of stillbirth and early neonatal death, compared to live births surviving more than 7 days, after adjusting for maternal age, household wealth, and parity. Total 8,806 singleton births among female residents under 35 years at birth, Baliakandi sub-district, Bangladesh, September 2017 to August 2019.

|  | **Stillbirth**  **Adjusted odds ratio (95% CI)** | **Early neonatal**  **Adjusted odds ratio (95% CI)** |
| --- | --- | --- |
| Maternal age at delivery (years) |  |  |
| 13 to 15 | 1.77 (0.92-3.38) | 1.03 (0.44-2.39) |
| 16 to 17 | 1.01 (0.61-1.67) | 1.28 (0.80-2.04) |
| 18 to 34 | Ref | Ref |
| Parity |  |  |
| 0 | 1.51 (1.07-2.14) | 1.68 (1.17-2.43) |
| 1 | Ref | Ref |
| 2 | 1.10 (0.69-1.77) | 1.41 (0.88-2.26) |
| 3+ | 1.63 (0.86-3.06) | 1.93 (1.02-3.67) |
| Household wealth quintile |  |  |
| Highest | 0.53 (0.33-0.86) | 0.84 (0.53-1.33) |
| High | 0.83 (0.55-1.26) | 0.72 (0.45-1.16) |
| Middle | 0.80 (0.52-1.22) | 0.92 (0.59-1.45) |
| Low | 0.75 (0.49-1.17) | 1.04 (0.66-1.62) |
| Lowest | Ref | Ref |
