## Supplementary figure 1 for "Persistence of child marriage in rural Bangladesh and impact on maternal and perinatal health: findings from a health and demographic surveillance system"

### Supplementary figure 1. Baliakandi Health and Demographic Surveillance System data and analyses used to estimate proportion of marriage to female children and time between first marriage and pregnancy in Baliakandi, Bangladesh.

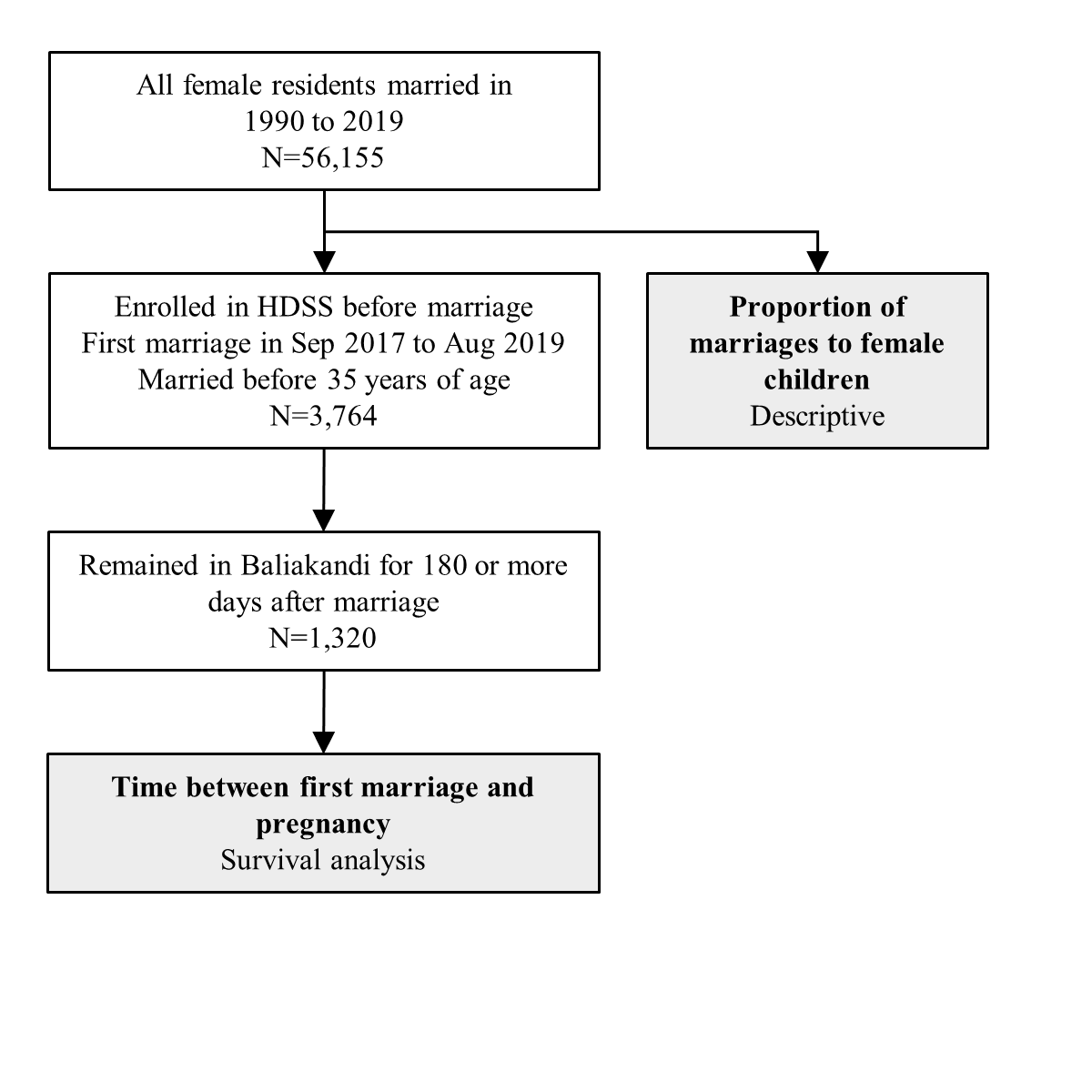
