## Supplementary figure 2 for "Persistence of child marriage in rural Bangladesh and impact on maternal and perinatal health: findings from a health and demographic surveillance system"

### Supplementary figure 2. Baliakandi Health and Demographic Surveillance System data and analyses used to examine relation between maternal age at birth, complications during delivery, and perinatal mortality in Baliakandi, Bangladesh

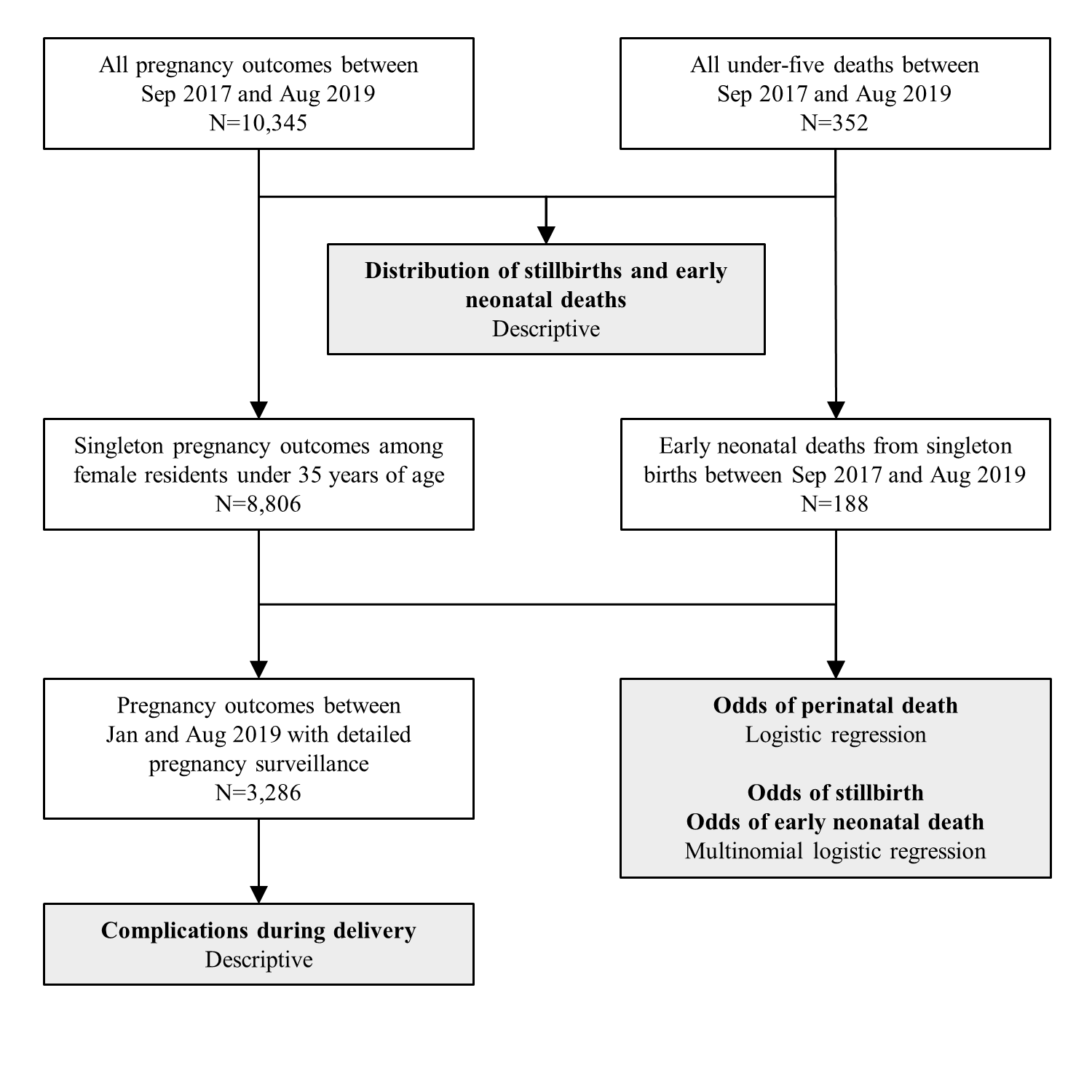
