## Supplementary table 1 for "Persistence of child marriage in rural Bangladesh and impact on maternal and perinatal health: findings from a health and demographic surveillance system"

### Supplementary table 1. Characteristics of female residents under 35 years of age who had first marriages, Baliakandi sub-district, Bangladesh, September 2017 to August 2019.

| **Characteristic** | **All**  **N=3,764**  **n (%)** | **Exited Baliakandi within 180 days of marriage**  **N=1,940**  **n (%)** | **Stayed in Baliakandi**  **N=1,824**  **n (%)** |
| --- | --- | --- | --- |
| Age at marriage (years) |  |  |  |
| 10 to 15 | 1077 (29%) | 499 (26%) | 578 (32%) |
| 16 to 17 | 1150 (31%) | 591 (30%) | 559 (31%) |
| 18 to 34 | 1537 (41%) | 850 (44%) | 687 (38%) |
| Household wealth quintile |  |  |  |
| Highest | 733 (19%) | 330 (17%) | 403 (22%) |
| High | 767 (20%) | 379 (20%) | 388 (21%) |
| Middle | 770 (20%) | 384 (20%) | 386 (21%) |
| Low | 759 (20%) | 414 (21%) | 345 (19%) |
| Lowest | 734 (20%) | 433 (22%) | 301 (17%) |
