## Supplementary table 2 for "Persistence of child marriage in rural Bangladesh and impact on maternal and perinatal health: findings from a health and demographic surveillance system"

### Supplementary table 2. Cumulative incidence of pregnancies after first marriage among 786 newly married female residents who remained in Baliakandi for at least 365 days after marriage, Baliakandi, Bangladesh, September 2017 to August 2019.

| **Age at marriage (years)** | **Cumulative incidence of pregnancies at 365 days after marriage (95% CI)** |
| --- | --- |
| 12-15 | 53% (46-59%) |
| 16-17 | 58% (50-64%) |
| 18-34 | 50% (46-55%) |
